# Variability in self-reported depression symptomology and associated behavioral markers in digital phenotyping

**DOI:** 10.1101/2025.03.26.25324604

**Authors:** Arsi Ikäheimonen, Nguyen Luong, Ilya Baryshnikov, Ti John, Annasofia Martikkala, Erkki Isometsä, Talayeh Aledavood

## Abstract

Digital phenotyping studies using smartphone-sensed data have identified several behavioral markers associated with depression. However, the generalizability of these markers is constrained by multiple factors, including variability in depressive symptoms and associated behaviors, both between and within individuals over time. This study examines heterogeneity in depression and aims to identify behavioral markers indicative of depression in smartphone-sensed data collected from participants diagnosed with depression. We analyzed smartphone-sensed behavioral data from 62 patients with major depressive episodes across three subgroups: major depressive disorder (MDD, n=41), borderline personality disorder (BPD, n=12), and bipolar disorder (BD, n=9). Depression symptoms were assessed with the 9-item Patient Health Questionnaire (PHQ-9). Symptoms varied between subgroups and across severity levels. Association analysis revealed variability in correlations between depression severity and behavioral markers, both between participants and over time. Multilevel modeling revealed demographic predictors: employment status (β = −4.79, 95% CI = [−7.65, −1.80], P = .004) and age (β = −0.12, 95% CI = [−0.25, −0.00], P = .050) and lower nighttime movement (β = −0.79, 95% CI = [−1.29, −0.29], P = .024), as behavioral markers of depression.

## Introduction

Depressive disorders rank among the most prevalent mental disorders and are among the most burdensome health conditions worldwide [1]. In 2021, an estimated 330 million people worldwide were living with depressive disorders, affecting approximately 4.4% of the global population [2]. Clinical assessment of depression relies on structural interviews, observer scales, and questionnaires [3], which are prone to biases such as recall bias [4]. Additionally, the heterogeneous nature of depressive symptoms [5] adds further complexity to diagnosis.

The ubiquity of smartphones and other personal smart devices has inspired research to use data from these devices to detect and predict depression symptoms objectively. Digital phenotyping is a field of research that utilizes personal digital devices to measure human behavior and physiology unobtrusively and continuously in real-life settings [6]. Collecting and analyzing data on individuals’ behavior using these devices can provide new information about mental health disorders, including depression and behavioral patterns associated with them [7]. Ideally, the knowledge produced by digital phenotyping research could be applied in practice as new methods and tools for symptom monitoring and prediction, supplementing current means of mental healthcare. Furthermore, these methods could help design early warning systems, which can be integrated with interventions for symptom worsening [8].

Research on digital phenotyping of depression has mainly focused on identifying associations between smartphone-sensed behavior and depression symptoms, predicting depression symptom severity and diagnostic classification, and forecasting future mood [9]. The studies typically rely on behavioral data representing participants’ physical activity, mobility, sociability, sleep, and smartphone usage patterns [10]. Although these approaches have achieved promising results, the reported associations between behavior and depression vary across studies in both statistical significance and direction (positive or negative) (see, e.g., [10,11]), implying that the results may not generalize well across different samples and contexts. The differences may arise partly from heterogeneity in study objectives, samples, used methodologies, and reporting practices [10], device usage differences between the study participants [12], but also from the differences in depressive symptoms between participants [13] and temporal variability of symptoms within the participants [14–18]. The heterogeneity in depression symptoms and related observable behavior characterizes the depressive disorder; for example, both insomnia and hypersomnia, decrease or increase in appetite, are regarded as depression symptoms [19]. Further, research has identified heterogeneous temporal characteristics of individual depression symptoms [20] and variability in long-term trajectories of depression severity and stability [21].

This study aims to explore variability in depression symptoms and related behavioral markers as measured through digital phenotyping. Moreover, it assesses the underexplored contribution of between- and within-participant effects in behavior-depression relationships, aiming to identify markers associated with depression. The distinction between these effects is important for evaluating how both long-term, between-participant differences in behavior and short-term fluctuations within participants’ behavior reflect depression severity. We investigated variability from two angles. The first angle looked at differences in self-reported depression symptoms through answers to questionnaires. The second angle focused on the smartphone-sensed behavioral markers and their associations with depression severity. The first angle was represented by our first study objective, where the questionnaire answers were compared at the symptom level, across diagnostic groups, and by severity level. The second angle led to our second and third study objectives. The second objective was to explore correlations between behavioral markers and depression across the study population, at the participant level, and over time. The third objective was to assess both within- and between-participant variability in behavioral features using multilevel modeling and identify behavioral markers indicative of depression while controlling for participants’ background and contextual factors.

The study analyzed smartphone-collected behavioral data collected from outpatients diagnosed with depression. Self-reported scores from the 9-item Patient Health Questionnaire (PHQ-9)[22] were used to assess the severity of depression. We extracted features from smartphone data, reflecting daily behavioral aspects: *physical activity*, *mobility*, *sociability*, *phone usage*, and *sleep*. These features served as proxies for behavioral patterns, possibly associated with depression.

Exploratory analysis found heterogeneity in depressive symptom representations between the patient groups and in behavior-depression associations across the study population. The association exhibited variability in strength and direction (positive and negative) over time within the participants. Furthermore, multilevel modeling demonstrated that the variability over time in behavior within individual participants was more predictive of depression severity than the behavioral differences between participants. These results suggest that the predictive power of smartphone-sensed behavioral markers may be limited when simple, unimodal linear models are applied at the population level (assuming that the same behavioral markers have similar associations with depression across participants). On the contrary, results indicate that modeling should focus on the behavioral changes over time within the participants and on individual symptoms for more accurate depression monitoring and prediction.

## Results

### PHQ-9 questionnaire analysis

Our first objective was to assess the variability in depression symptoms through PHQ-9 answers at item (symptom), diagnostic group, and severity levels. The summary statistics showed that the MDD patients had, on average, the lowest average depression level (mean=11.6, SD = 6.0), while BPD had the highest average (mean=14.9, SD=4.8). At the study population level, moderate depression severity (PHQ-9 scores from 10 to 14) was the most frequent (27.5%). For a group-level summary of depression severity, refer to Table S1 in Multimedia Appendix 1. Considering PHQ-9 score variability, the BD group showed a wider distribution than others, reflecting higher within-group differences in depression variability. Nonetheless, the Kruskal-Wallis test [23] indicated that these differences were not statistically significant. Figure 1 shows the distribution of PHQ-9 mean scores and standard deviations for each group.

**Figure 1.**
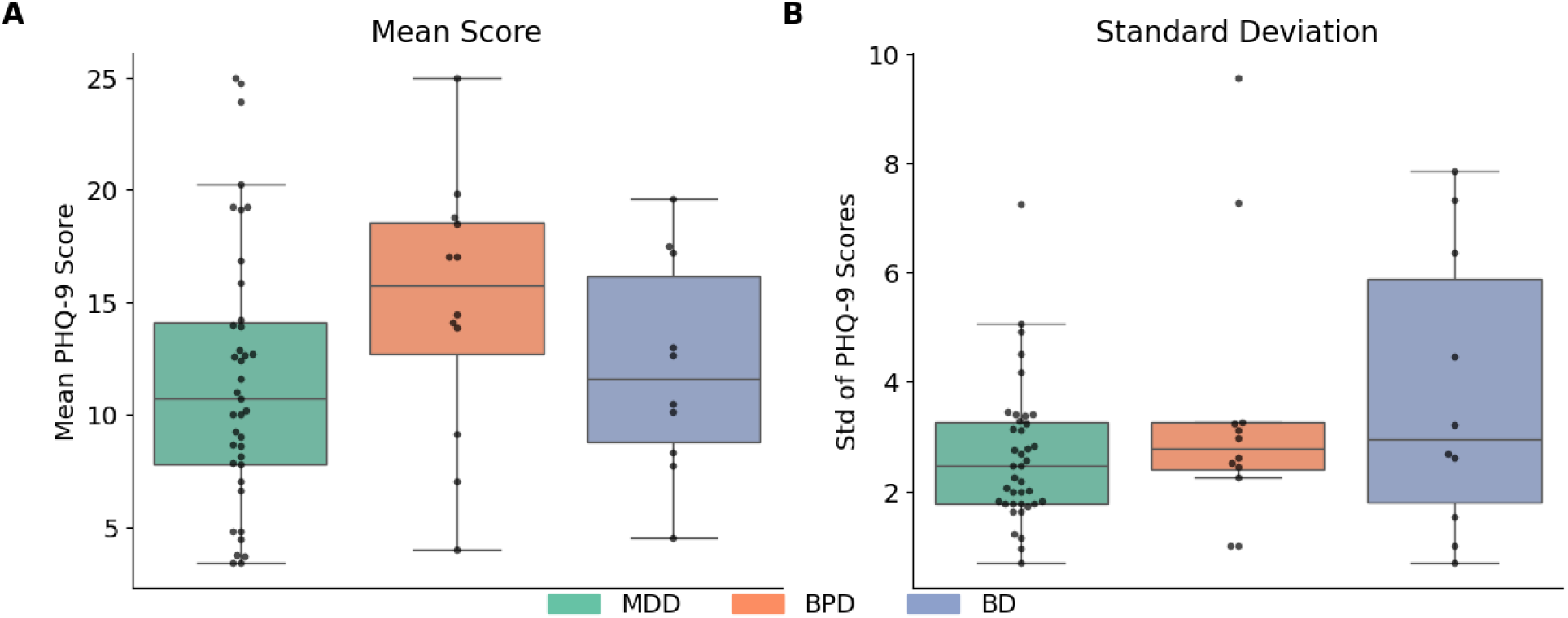
PHQ-9 score group-level summary statistics. The scores were first averaged within participants and then visualized. A) Boxplots of group-level mean scores: MDD (μ=11.65, *σ*=5.96), BPD (μ=14.89, *σ*=5.87), and BD (μ=12.12, *σ*=4.83), B) Boxplots of standard deviations of scores: MDD (μ=2.65, *σ*=1.28), BPD (μ=3.44, *σ*=2.49), and BD (μ=3.78, *σ*=2.61). A wider interquartile range for BD suggests greater fluctuations in depression severity among BD patients compared to others.

An exploratory comparison of the PHQ-9 item distributions at different depression severity levels exhibited differences across the patient groups. However, conducting the Kruskall-Wallis test indicated that these differences were not statistically significant. At the lowest depression level, items 3 (trouble with sleep) and 4 (tiredness or low energy) are more pronounced in the BPD group, while item 7 (trouble concentrating) has higher scores for the BD group than others. At the mild and moderate depression levels, the items are more evenly distributed across all groups. At the moderately severe level, item 9 (suicidal ideation) scores higher in the BD group, while item 1 (anhedonia) is pronounced in the MDD group, and the BPD group also exhibits higher scores for item 6 (low self-worth). Finally, at the highest severity level, the BD group differs from the BPD group, particularly in items 8 (psychomotor retardation) and 9 (suicidal ideation). Figure 2 illustrates these differences in distributions across groups and depression severity levels. For detailed results, refer to Appendix 1 Table S2, and Table S3 for a summary of PHQ-9 questions.

**Figure 2.**
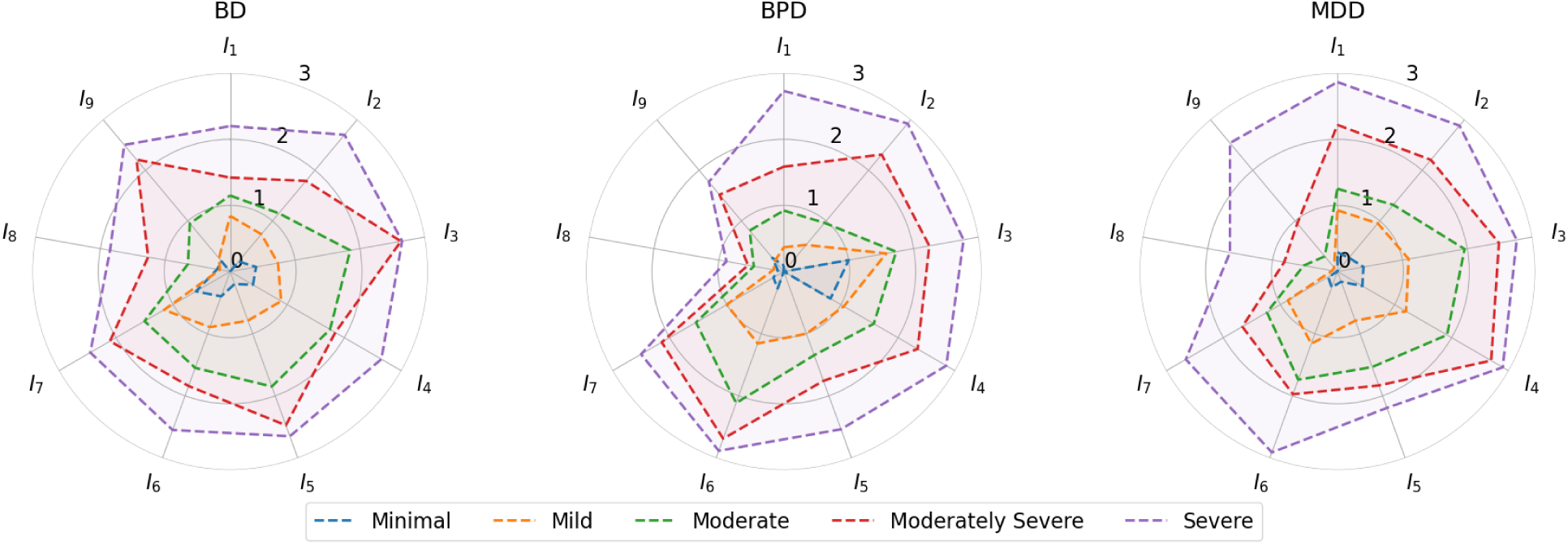
Average PHQ-9 item scores by diagnosis group and depression severity. For visualization purposes, we treated PHQ-9 items (ordinal) as continuous variables. The radar charts show the average of PHQ-9 item scores across different depression severity levels for BD, BPD, and MDD groups. Letters from I_1_ to I_9_ denote the items, and different colors represent severity levels. The scale, ranging from 0 (center) to 3 (outer circle), denotes item values.

Further exploration of the item score distributions across depression severity levels revealed that, on average, PHQ-9 items do not increase at the same rate or linearly with the total score. Items 3 (trouble with sleep) and 4 (tiredness or low energy) constantly contributed to the total score across all the severity levels. Also, item 6 (low self-worth) had high scores across the severity levels. On the other hand, item 8 (psychomotor retardation) stayed relatively low across all depression severity levels. Items 3, 4, 5 (poor appetite or overeating), and 6 showed the highest relative changes, transiting between mild depression and moderate depression. These changes suggest that these items (and corresponding symptoms) might be particularly informative for detecting depression severity above the clinical threshold (a PHQ-9 score of 10). Further, item 9 (suicidal ideation) only emerged at higher depression levels. Figure 3 illustrates the PHQ-9 item distributions across the severity levels.

**Figure 3.**
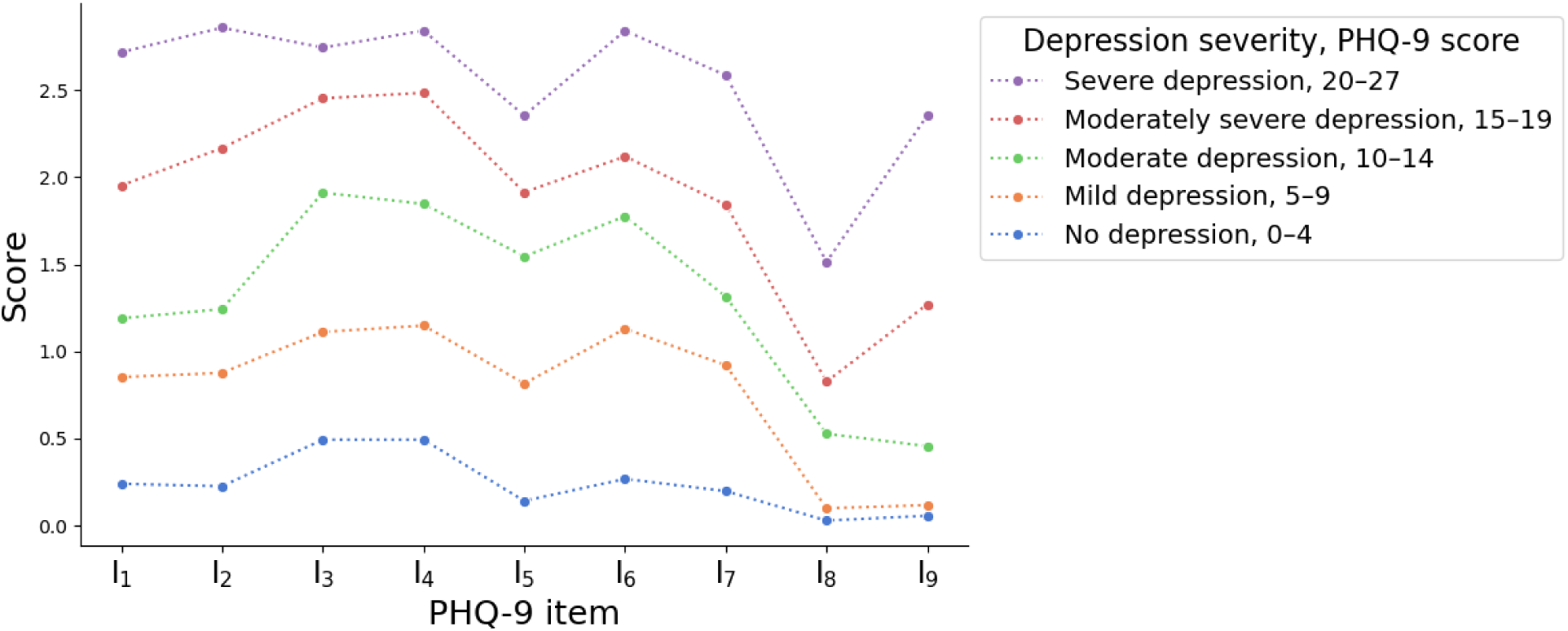
PHQ-9 item average score distributions across depression severity levels, combining the data from all patient groups. For exploratory analysis purposes, we treated the (ordinal) items as continuous variables and visualized the distribution shapes. The comparison shows that the items neither increase at the same rate nor in a linear manner with total depression severity. Notably, items 8 and 9 have higher values only at higher severity levels. Additionally, comparing the mild (scores from 5 to 9) and moderate (scores from 10 to 14) levels of depression severity, items 3 to 6 exhibit the highest differences.

### Association analysis

To address the second objective, we examined the associations between behavioral markers and depression severity. We conducted an exploratory correlation analysis, calculating pairwise Kendall rank correlation coefficients between the behavioral features and PHQ-9 scores by pooling data from participants, and at the individual participant level, thus comparing general, study population-level associations and individual participant-level variability. The results revealed that pooling the participants, all behavioral data features had weak to moderate correlations (Kendall rank correlation coefficient between −0.3 and 0.3) with PHQ-9 depression scores. Nonetheless, these correlations were not significant after controlling for false positives with false discovery rate (FDR) correction at a significance level of *α* = .05. On the other hand, inspecting the correlations at the individual participant level resulted in a broader range of coefficients, summarized in Table 1.

**Table 1.**
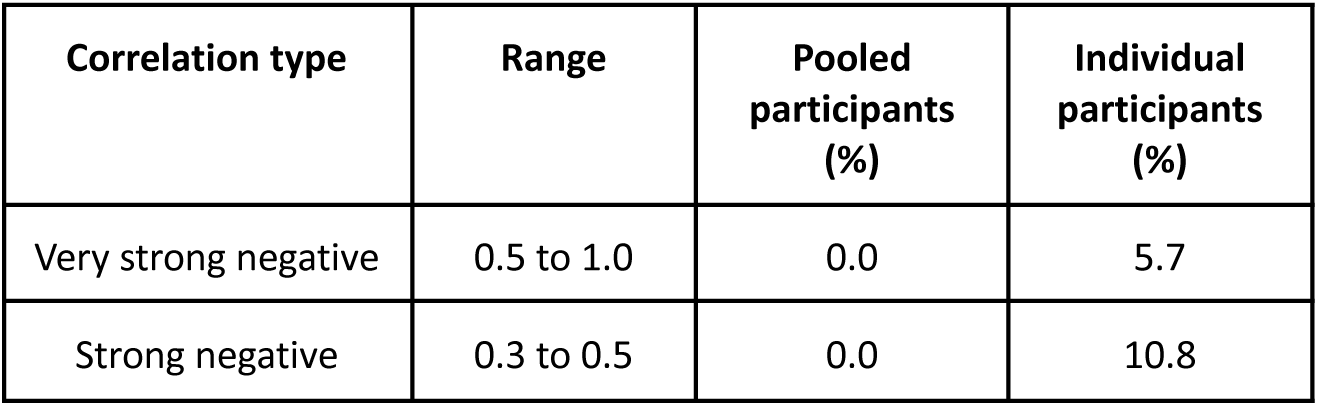
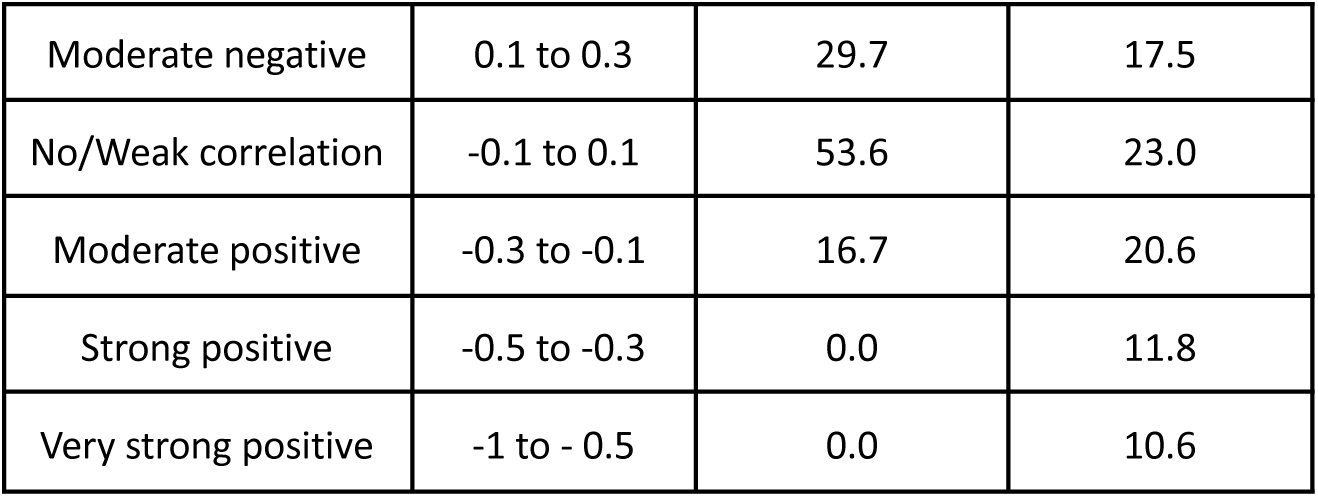
Association analysis summary, the proportion of correlation types across the behavioral features. The correlation was measured by Kendall rank correlation coefficients, calculated by pooling the participants and at the participant level. In general, participant-level coefficients exhibited broader distributions than pooled-level coefficients.

Further, we examined how the associations varied over time within the participants with exploratory rolling-window correlation analysis. We counted the proportion of users and features exhibiting both positive and negative correlations during the data collection period across different window sizes. The results indicated that correlation directions (positive or negative) might change over time. In general, the proportions of mixed correlation, reflecting that participants had both positive and negative correlations, are higher with shorter windows. These findings suggest that the associations are not static and should be considered dynamic in modeling tasks. Appendix 1 Figure S1 summarizes the average proportion of features exhibiting both positive and negative correlations across participants.

### Multilevel modeling

To address the third objective, we used multilevel modeling to examine both within- and between-participant variability in behavior and to identify markers indicative of depression. We employed a stepwise modeling strategy, starting with a baseline model that included only the random intercepts for each participant and subsequently adding background information, between- and within-effects as predictors. The baseline model, including only random intercepts, explained 72.0% of the total variance. The first model, which included demographic and contextual information, increased the conditional R^2^-score (accounting for both fixed and random effects) to 0.737 and the marginal R^2^-score (reflecting only fixed effects) to 0.162. The second model, adding time-varying within-participant features, further increased the conditional R^2^ to 0.743 and the marginal R^2^-score to 0.172. Finally, the complete model combines background, contextual, and within- and between-participant behavioral features, resulting in a conditional R^2^-score of 0.797 and a marginal R^2^-score of 0.207.

The results from multilevel models showed that the intraclass correlation coefficient (ICC) was relatively high across models (ranging from 0.69 to 0.73), indicating that a substantial proportion of the variability in self-reported depression severity was attributed to the between-participant differences. Model comparisons with ANOVA revealed that adding within-participant behavioral features (Model 2) improved model fit over the baseline model (Model 1) (χ² = 26.22,, df = 16, P = .05; AIC = 3613.76, BIC = 3721.02). However, incorporating between-participant features (Model 3) only led to marginal improvement over Model 2 (χ² = 54.78, df = 16, P < .001; AIC = 3590.9, BIC = 3769.74). Therefore, Model 2 represents the most parsimonious model.

In Model 2, employment status (β = −4.79, 95% CI: −7.65 to −1.80, P = .004) and age (β = −0.12, 95% CI: −0.25 to −0.00, P = .050) emerged as significant demographic predictors of depression severity, with unemployment and younger age linked to higher symptom levels. Among behavioral features, higher nighttime movement (β = −0.35, 95% CI: −0.63 to −0.06, P = .024) was linked to reduced symptom severity. Notably, when taking into account between-participant effect, outgoing SMS count emerge as a significant predictors. The effect is positive at between-participant level (β = 2.20, 95% CI: 1.12 to 3.32, P = <.001) but negative at within-participant level (β = -.56, 95% CI: −1.00 to −0.17, P = .004). Table 2 shows the condensed model summaries, displaying only the features with P-values below 0.05. For the detailed results, refer to Table S4 in Multimedia Appendix 1.

**Table 2.**
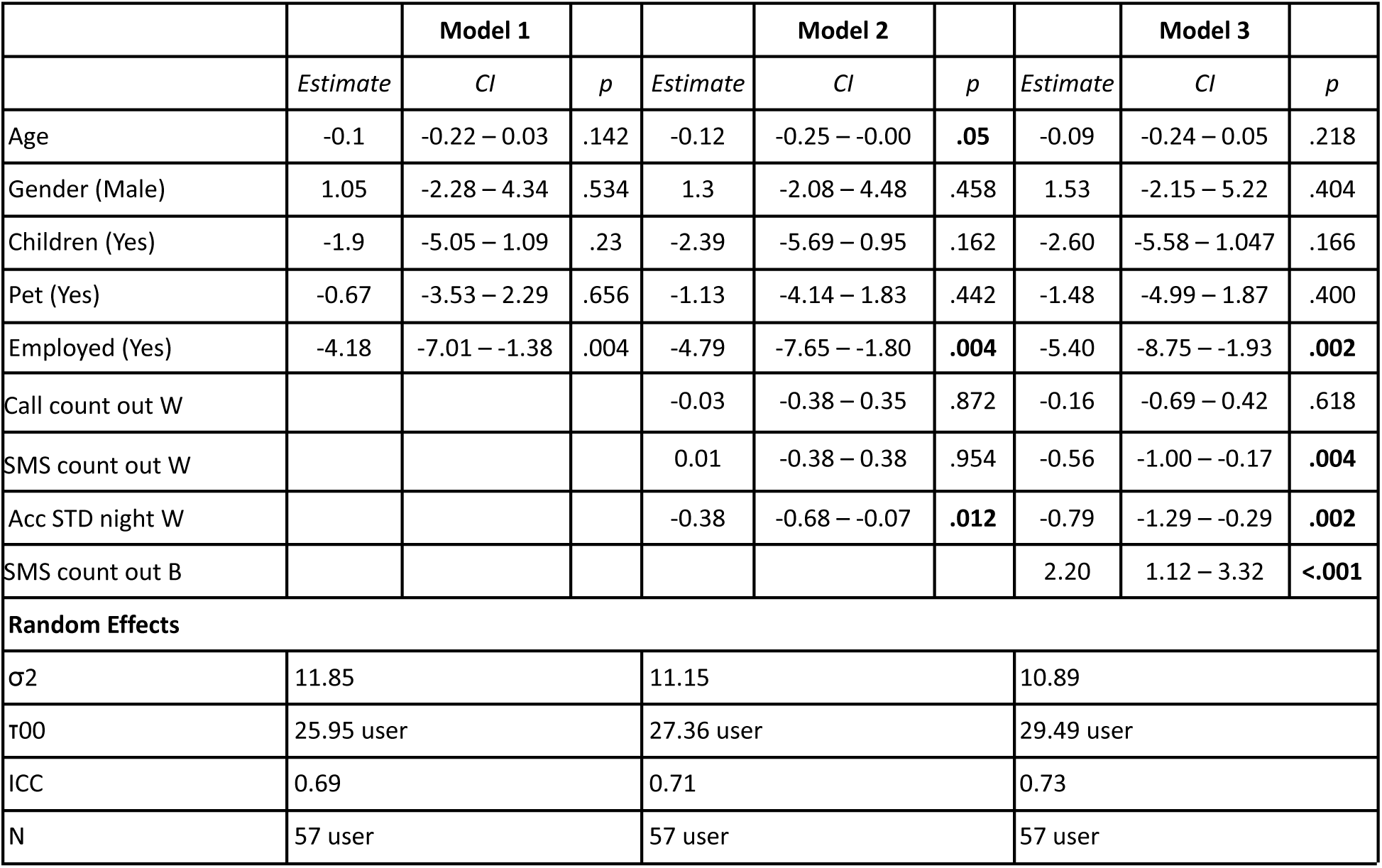
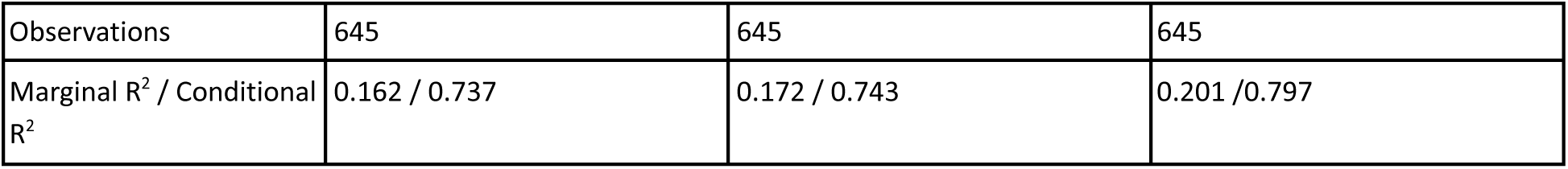
Multilevel model summaries. For brevity, the table shows only the statistically significant behavioral predictors. Model 2 is the best-fitting and most parsimonious, showing within-participant behavioral features: nighttime accelerometer variability with lower severity. Additionally, higher age and employed status are associated with lower depression severity. W denotes the within-participant effect, σ2 residual variance, τ00 intercept variance, and ICC intra-class correlation coefficient. Model 1 includes background demographics and contextual predictors, with random intercepts for each participant. Model 2 adds within-participant effects (features) into model 1 Model 3 adds between-participant effects (features) into model 2.

|  |  | Model 1 |  |  | Model 2 |  |  | Model 3 |  |
| --- | --- | --- | --- | --- | --- | --- | --- | --- | --- |
|  | *Estimate* | *CI* | *p* | *Estimate* | *CI* | *p* | *Estimate* | *CI* | *p* |
| Age | -0.1 | -0.22 – 0.03 | .142 | -0.12 | -0.25 – -0.00 | .05 | -0.09 | -0.24 – 0.05 | .218 |
| Gender (Male) | 1.05 | -2.28 – 4.34 | .534 | 1.3 | -2.08 – 4.48 | .458 | 1.53 | -2.15 – 5.22 | .404 |
| Children (Yes) | -1.9 | -5.05 – 1.09 | .23 | -2.39 | -5.69 – 0.95 | .162 | -2.60 | -5.58 – 1.047 | .166 |
| Pet (Yes) | -0.67 | -3.53 – 2.29 | .656 | -1.13 | -4.14 – 1.83 | .442 | -1.48 | -4.99 – 1.87 | .400 |
| Employed (Yes) | -4.18 | -7.01 – -1.38 | .004 | -4.79 | -7.65 – -1.80 | .004 | -5.40 | -8.75 – -1.93 | .002 |
| Call count out W |  |  |  | -0.03 | -0.38 – 0.35 | .872 | -0.16 | -0.69 – 0.42 | .618 |
| SMS count out W |  |  |  | 0.01 | -0.38 – 0.38 | .954 | -0.56 | -1.00 – -0.17 | .004 |
| Acc STD night W |  |  |  | -0.38 | -0.68 – -0.07 | .012 | -0.79 | -1.29 – -0.29 | .002 |
| SMS count out B |  |  |  |  |  |  | 2.20 | 1.12 – 3.32 | <.001 |
| Random Effects |  |  |  |  |  |  |  |  |  |
| σ2 | 11.85 |  |  | 11.15 |  |  | 10.89 |  |  |
| τ00 | 25.95 user |  |  | 27.36 user |  |  | 29.49 user |  |  |
| ICC | 0.69 |  |  | 0.71 |  |  | 0.73 |  |  |
| N | 57 user |  |  | 57 user |  |  | 57 user |  |  |

|  |  |  |  |
| --- | --- | --- | --- |
| Observations | 645 | 645 | 645 |
| Marginal R<sup>2</sup> / Conditional R<sup>2</sup> | 0.162 / 0.737 | 0.172 / 0.743 | 0.201 / 0.797 |

## Discussion

This study investigated the variability in self-reported symptoms of depression and associated smartphone-sensed behaviors. We observed variability in PHQ-9 items between the patient groups and across different levels of depression severity. Associations between behavioral markers and depression severity varied considerably between the study participants and over time within participants. The multilevel model results indicate that approximately 70% of the variability in self-assessed depression is attributable to between-participants differences in depression severity. Adding demographic background, context, and time-varying within-participant behavioral features improves the model’s explanatory power by up to 76.3%. Multilevel modeling identified that demographic factors, age and employment status, and within-participant behavioral markers of daily outgoing SMS count and physical nighttime activity are associated with depression severity.

Inspection of PHQ-9 item distributions revealed differences in the representation of depression across the diagnostic subgroups. The differences suggest that accommodating individual differences in depression manifestations at the symptom level and modeling each participant individually or pooling together participants with similar symptoms might benefit the modeling accuracy. Additionally, these differences in symptom profiles may serve as proof of the principle that distinct subgroups exist within depressive disorder; a unique symptom profile may characterize each subgroup. However, as the subgroups in this study were small, caution should be exercised when generalizing such findings.

Further, we observed the highest difference in PHQ-9 score distributions between mild and moderate depression on items 3 (trouble with sleep), 4 (tiredness or low energy), 5 (poor appetite or overeating), and 6 (low self-worth). These differences suggest that the transition from lower depression into moderate level may be identifiable through monitoring behavior related to these items, e.g., sleep disturbances, daily activity and mobility, and sociability. Additionally, we noticed that item 8 (psychomotor retardation) exhibits higher scores only at higher depression severity levels, aligning with clinical knowledge that psychomotor retardation typically occurs only in severe depression and is an important characteristic of the DSM-5 melancholic specifier or subtype of major depressive episodes [19,24]. Similarly, item 9 (suicidal ideation) gets higher scores only at higher depression levels. This finding is consistent with other studies and is a clinically important point - the alleviation of depression results in the alleviation of suicidal thoughts [25].

Correlation analysis results indicated that most behavioral markers had weak correlations with PHQ-9 depression scores at the population level. In contrast, at the individual participant level, the correlations exhibited a wider range of both positive and negative values, which may cancel each other out when aggregated at the population level. Notably, the number of data points provided by each participant varies. Thus, some of the higher correlations might stem from the relatively low observation count by change. On the other hand, the low variability in PHQ-9 scores could explain the low correlations for some participants. Likewise, considering application usage-related markers, low correlations may occur because some participants use smartphone applications infrequently. Finally, the complementary rolling-window correlation analysis explored the within-participant variability in time, revealing periods with varying correlation strengths and directions. This observation suggests that the relationship between behavioral markers and depression severity should be considered time-dependent rather than static, aligning with existing literature that emphasizes the need for models that can accommodate the time-varying dynamics of the depression-behavior relationship [14,16].

The multilevel modeling results regarding the explanatory power of random and fixed effects align with other studies using hierarchical linear modeling (e.g., [26–28]). Adding behavioral features increased the marginal R^2^ -score by 2.1%, indicating modest explanatory power of passively sensed, time-varying participant-level behavioral features. Further, the significant portion of PHQ-9 variance is explained by between-participant variability in depression severity. These findings suggest focusing on within-participant effects in behavior and accounting for participant’s depression severity history in detection and future prediction.

The validity and generalizability of this study’s findings are primarily limited by the amount of available data; the sample size, participants dropping out of the study, and missing intermediate observations (the dropout rate and data missingness is more thoroughly discussed in our previous work [29,30]). Limited yet high dimensional data is a common issue across studies involving personal digital devices for data collection [31,32]. While the quantity of passively collected raw data is relatively high in this study, more bi-weekly sampled active data for each participant would have benefited the analysis, leading to higher statistical power and making modeling less prone to overfitting. Also, combining data sets across different studies may provide a feasible approach to obtaining larger and more heterogeneous samples.

Additionally, the temporal nature of the data might bias the analysis results. Used feature standardization and imputation techniques assume stationarity of variance over time, which might not hold with behavior in participants with mood disorders. These issues could be mitigated by considering time-aware standardization and thoroughly analyzing the nature of missing observations (whether they are missing completely at random, at random, or not at random) and thorough comparison between different imputation techniques, such as multiple imputation using chained equations (MICE) [33].

It is also worth exploring whether the changes in behavior are more common in moderate to severe depression. If sensitive digital phenotyping measures for the milder range of depression are to be explored, they probably must tackle more subjective and internal aspects of depression. Achieving this might require measures that are sensitive to the negative emotional and self-referential cognitive biases in depression, such as changes in speech prosody or the linguistic content of written texts (for example, in emails and social media platforms), consumed media content, or facial expressions.

Due to the limited explanatory power of linear modeling, we recommend using multiple features in future modeling, exploring feature interactions and non-linear relationships in the data with modeling techniques, such as gradient-boosting trees, and personalizing the models. Previous research has addressed the problem of between-participant variability through personalized modeling that adapts to the differences between participants. The models can be personalized, for example, by using a participant-specific (idiographic) approach (e.g., [28]), personalizing the depression symptom profiles (e.g., [34]), hierarchical modeling (e.g., [35,36]), symptom similarity-based grouping (e.g.,[37]), combining the training data from participants with similar behavioral feature distributions (e.g., [38]), or collaborative filtering modeling (e.g.,[39]).

This study exemplifies how self-reported depressive symptoms and associated mobile-sensed behaviors vary between and within patients clinically diagnosed as experiencing major depressive episodes. Our multilevel modeling revealed that a substantial portion of depression variability is due to individual differences. In summary, these findings suggest that smartphone-sensed behavioral markers may have limited clinical utility in monitoring depression using digital phenotyping within this study population. Hence, we propose that future research focus on improving depression prediction and forecasting accuracy by utilizing data from multiple sources using advanced techniques capable of modeling non-linear and time-varying relationships, feature interactions, and differences between participants. Through these results, we emphasize the importance of addressing and reporting heterogeneity within the study population, as it may provide critical information for future research design, identifying useful behavioral markers, and study sample size optimization with power analysis.

## Methods

### Setting

This study analyzed behavioral data from the Mobile Monitoring of Mood (MoMo-Mood) study [29], which includes continuously sensed smartphone behavioral data (passive data), self-reported responses to biweekly depression questionnaire (active data), and background demographics and contextual information. The study involved 164 participants across three patient groups, major depressive disorder (MDD, n=85), bipolar disorder (BD, n=21), borderline personality disorder (BPD, n=27), and a healthy control group (n=31).

Patients were voluntarily recruited from primary care and psychiatric outpatient treatment clinics in Finland, while healthy controls were recruited among university students and healthcare personnel. The patients were under treatment and diagnosed as having ongoing major depressive episodes with structured interviews, the Mini-International Neuropsychiatric Interview [40], and the Structured Clinical Interview for DSM-IV Axis II Personality Disorders [41]. The study used continuous enrolling, allowing the participants to enter or withdraw from the study at different times, with a recommended participation period of one year. All participants were informed about the study prior to enrolment, including the data collection and their option to leave the study at any time. Informed written consent was obtained from all the study participants. The participants were compensated with four movie tickets. The Helsinki and Uusimaa Hospital District’s Ethics Committee approved the study research protocols (including the data stream and collection platform security), and Helsinki and Uusimaa Hospital District Psychiatry granted a research permit (approval number § 125/2018). A written data security statement was authorized by the local research ethics committee and IT support. All research procedures followed the ethical standards of the Declaration of Helsinki. For more details, refer to earlier work [29,30,42].

### Dataset description

The passive data used in our analysis originated from various smartphone sensors and logs, collected using the AWARE app [43] and curated using the Niima platform [44]. The data encompasses five aspects of daily behavior: physical activity and mobility, sociability, phone interactions, and sleep. Behavioral features related to these aspects have been identified as being associated with depression severity in previous research ( e.g., for physical activity [45–47], mobility [48,49], sociability [50,51], phone interactions [52,53], and sleep [39,54]). We operationalized these behavioral aspects through specific features extracted from raw smartphone data, serving as proxies for real-world behaviors. The active data has participants’ self-reported PHQ-9 questionnaires, which assess the depression severity. The PHQ-9 is a tool used for screening for diagnosis and monitoring of depressive symptoms [55]. The PHQ-9 survey comprises nine items, each addressing the main symptoms of depression, such as having little interest or pleasure in doing things. According to the survey, patients were asked to respond based on their experience over the last two weeks, with each item rated from 0 (not at all) to 3 (nearly every day). The Background demographics include categorical variables, including participants’ age and sex, used as control variables. Contextual information includes details about full-time work, shift work, cohabitation status, the number of children, and whether the participants have pets. We selected these factors since they may affect participants’ daily behaviors.

### Data preparation and preprocessing

Of the 164 participants, we excluded the control participants from the analysis since they exhibited low levels and variability (mean = 1.2, std = 1.8) of self-measured depression [30]. Further, we excluded the participants who provided less than one month of passive data (equaling at least two biweekly PHQ-9 responses), resulting in a total of 62 (39%) participants from three different groups: (1) patients with BD (n=9), (2) patients with BPD (n=12), and (3) patients with MDD (n=41).

The raw passive data comprised accelerometer readings, application usage logs, battery level, GPS locations, phone call logs, SMS logs, and screen usage. We used the Niimpy behavioral analysis toolbox [56] to preprocess the raw sensor data and extract behavioral features. We extracted three types of features: *volume-based*, *temporal*, and *contextual features*. The volume-based features represent counts and durations of specific activities, such as smartphone screen usage time. Temporal features were derived by segmenting the volume-based features into 6-hour bins based on the time of day (12:00 AM-6:00 AM, 6:00 AM-12:00 PM, 12:00 PM-6:00 PM, and 6:00 PM-12:00 AM) for more precise information in daily rhythms, similar to our previous work [30]. Considering the behavioral aspects, physical activity was captured through phone accelerometer data, providing temporal information about movement intensity and variability. Mobility was assessed using GPS location data, which provided spatial (e.g., total distance traveled) and contextual information (e.g., daily time spent at home) on behavior. Sociability was measured by phone call counts, durations, and the number of messages sent and received. Phone interactions were inferred from application usage logs (e.g., application category, usage count, and duration), screen usage counts (on and off), usage duration, and battery charge level. Sleep duration and timing were approximated using the longest period of inactivity measured through screen activity data, following a process similar to that demonstrated in [29]. This method gives comparable results with other methodologies for measuring sleep in this population [57]. For further details, refer to Table S1 in Multimedia Appendix 1.

For active data, we used scores from individual items and assessed the depression severity with the total sum of item score, with the result ranging from 0 (no depression) to 27 (severe depression). Further, categorical background demographics and contextual variables were used as control variables. We used demographics, including age and gender, as well as contextual variables that might influence daily behavior: such as having children, owning a pet, and being employed. The variables were used in multilevel modeling, encoded using one-hot encoding.

Extracted temporal and volume-based behavioral features were aligned with biweekly depression questionnaires and aggregated by calculating the arithmetic mean over the 14 days preceding the days the PHQ-9 measurement took place. Location data features (including distance-based and significant places-based features) were calculated directly over 14-day windows, thus requiring no averaging. Further, we pre-filtered redundant features based on prior knowledge (for example, screen off count, since it reflects the same information as screen on count). The resulting data comprised 671 biweekly observations covering 224 behavioral features, two background demographics, five contextual variables, and one external variable. Figure 4 shows the details of data preprocessing and aggregation schematics.

**Figure 4.**
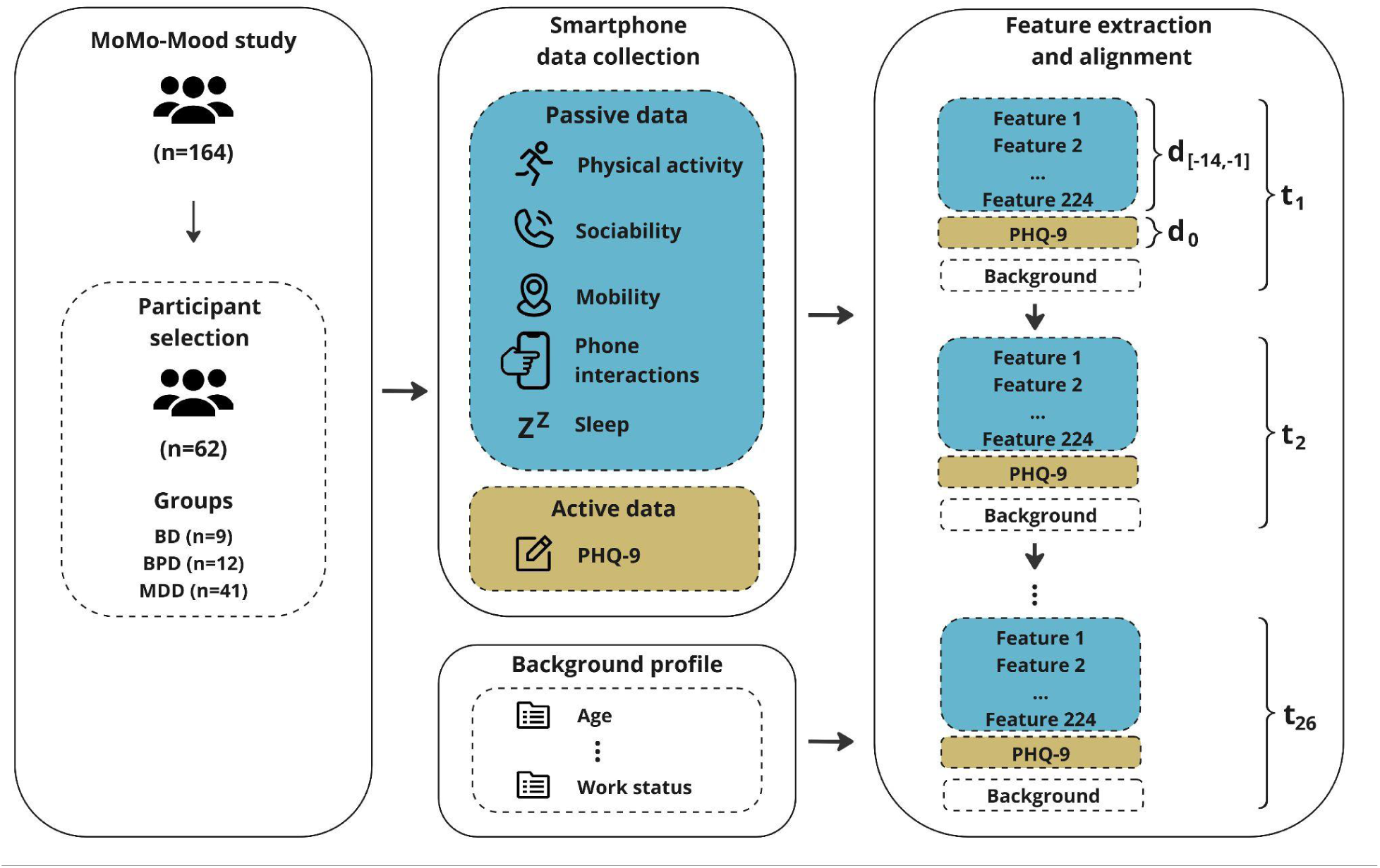
Participant selection, data collection, and processing schema. For this study, we selected participants from three patient groups (BD, BPD, and MDD) who answered at least two bi-weekly (14-day) PHQ-9 questionnaires and provided passive data for 14 days prior to each PHQ-9 answer. Smartphone-collected data encompassed: 1) passive data, consisting of raw smartphone-sensed data, including accelerometer readings, phone call and SMS logs, app and screen usage logs, battery level logs, and GPS location data; 2) active data, consisting of PHQ-9 questionnaires; and 3) demographic information, such as participants’ age and work status. The merged datapoints are denoted by t_1_ to t_26_, and day d_0_ represents the day the PHQ-9 was reported, and d_-14_ to d_-1_ the preceding 14-day period passive data was collected. Data collection lasted up to one year.

For the first study objective, to assess how depressive symptoms differ between patient groups, we used the raw values from the PHQ-9 questionnaire items, removing missing or incomplete records. For the second objective, behavior–depression association analysis, we used aligned passive (behavioral features) and active (self-assessed PHQ-9 scores) data, removing observations where either the feature or questionnaire was missing. For the third objective, we imputed the missing intermittent passive data observations. Behaviors that do not tend to occur daily, such as sociability, were imputed with zeros. Within-participant median imputation was used for the remaining variables. Further, to examine within- and between-participant effects of behavior using multilevel modeling, we disaggregated passive data features using participant-mean and grand-mean centering [58]. The within-participant components reflect fluctuations relative to an individual’s average and their relationship to the severity of depression. Contrarily, between-participant effects reflect how differences in average behavior relate to depression severity (e.g., whether participants sleeping longer on average have higher or lower depression levels). For within-participant centering, we centered each variable around the participant’s mean to account for variability over time. Second, for between-participant centering, the variables were averaged within participants and then centered across the study population’s mean. This centering approach enables the detection of both individual-level behavioral fluctuations and broader population-level patterns.

### PHQ-9 questionnaire analysis

To assess the heterogeneity in self-reported depression, we conducted a descriptive analysis of PHQ-9 scores, applied the Kruskal-Wallis test [23] to evaluate group-wise differences in score means and standard deviations, and performed an exploratory analysis for each item across the groups and different depression severity levels. Additionally, we utilized exploratory data analysis to examine the scores across the groups and severity levels

### Association analysis

To investigate the associations between behavioral features and self-reported depression among participants, we conducted exploratory correlation analysis, calculating pair-wise Kendall rank correlation coefficients between the behavioral features and PHQ-9 scores. We selected Kendall rank correlation since it does not require the compared variables to be normally distributed or linearly related, and it can be used with ordinal data. For analysis, we used a function implemented in the *SciPy* library [59]. We employed two approaches: pooling data from all participants and calculating the coefficients for each participant individually. Due to the multiple comparisons, we applied the FDR correction with the Benjamini-Hochberg procedure [60] to control Type I errors. Additionally, we examined whether the associations between behavioral features and depression symptoms varied within participants over time by conducting an exploratory rolling-window correlation analysis. We tested various window sizes, ranging from 28 days to one year. Within each window, we computed the Kendall rank correlation coefficient, shifting the window forward by 14 days to calculate successive correlations. We excluded participants who did not have enough data to fill at least one complete window for a given window size (and subsequent longer window sizes) from the analysis. For each window size, we used 1,000 bootstrapping samples to generate more robust distributions of correlation coefficients.

### Multilevel modeling

To assess the within- and between-participant effects of behavioral features on PHQ-9 scores, we used multilevel modeling with random intercepts to account for the data’s hierarchical structure. Independent variables consisted of features representing behavioral aspects, including sleep, activity, mobility, sociability, and device usage. PHQ-9 scores were used as the dependent variable. Participants were used as grouping variables. Generalized variance inflation factor (GVIF) was used to assess multicollinearity, keeping only variables with GVIF < 10. We employed a stepwise modeling strategy. First, we started with a baseline model that included only random intercepts for each participant. Second, we included demographic and contextual variables. Next, within-participant variables were added to the model. Finally, between-participant variables were included. The *lme4* package [61] in R was used for fitting the models, and 95% confidence intervals were estimated using bootstrapping (1000 iterations) with *the sjPlot* package [62]. For model comparison, we used the likelihood ratio tests to assess explanatory power and ensure model parsimony. The model formulas and the final list of predictors are shown in Appendix 3.

## Data availability

The dataset analyzed in the current study is not publicly available, as the conditions of the granted research permit restrict its use in order to protect the participants’ privacy.

## Code availability

The code used for preprocessing the data is available on GitHub [63].

## Acknowledgments

The computational resources provided by the Aalto Science-IT project are gracefully acknowledged. The collection of data used in this manuscript was made possible with the help of Richard Darst, Jesper Ekelund, Roope Heikkilä, Joel Holmén, Kirsi Riihimäki, and Outi Saleva. We thank them for their valuable efforts at different stages. This research was supported by Wihuri Foundation through a personal research grant to Arsi Ikäheimonen.

## Contributions

**Arsi Ikäheimonen:** Conceptualization, Data analysis, Data preprocessing, Methodology, Study Design, Visualization, Writing – original draft, Writing – review & editing, **Nguyen Luong:** Data preprocessing, Methodology, Study Design, Multilevel-modeling, Writing – review & editing, **Ilya Baryshnikov:** Methodology, Data collection, **Ti John:** Methodology, Study Design, Writing – review & editing, **Annasofia Martikkala:** Data collection, Writing – review & editing, **Talayeh Aledavood:** Conceptualization, Data collection, Methodology, Project administration, Supervision, Writing – review & editing. **Erkki Isometsä:** Conceptualization, Data collection, Project administration, Supervision. All authors reviewed the manuscript and approved the final version.

## Abbreviations

AIC: Akaike information criterion
ANOVA: Analysis of Variance
BD: Bipolar Disorder
BH: Benjamini-Hochberg
BIC: Bayesian information criterion
BPD: Borderline Personality Disorder
CI: Confidence Interval
FDR: False Discovery Rate
GPS: Global Positioning System
GVIF: Generalized Variance Inflation Factor
HUS: Helsinki and Uusimaa Hospital District
ICC: Intraclass Correlation Coefficient
MDD: Major Depressive Disorder
MICE: Multivariate Imputation by Chained Equations
PHQ-9: 9-item Patient Health Questionnaire
SMS: Short Message Service

## Multimedia Appendix 1

**Table S1.**
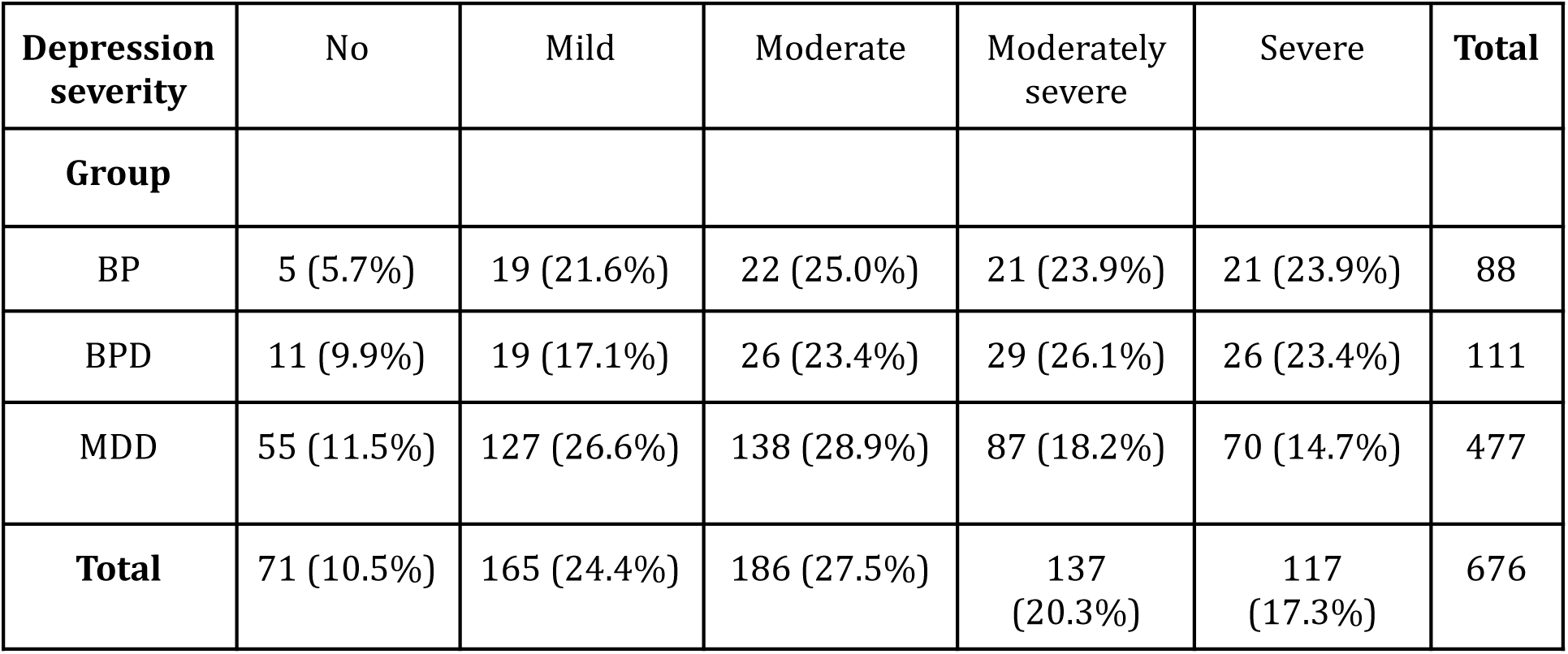
Depression severity levels by groups. The table shows the distribution of depression severity levels based on repeated measurements of PHQ-9, meaning that each user may contribute to multiple severity levels. The table shows that moderate depression severity (PHQ-9 scores from 10 to 15) is most frequent across the groups, and moderately severe and severe levels are more frequent among BD and BPD patients compared to MDD patients.

**Table S2.** *Figure 2* Median PHQ-9 item scores by diagnosis group and depression severity. The item scores range from 0 to 3.

|  | PHQ-9 item | 1 | 2 | 3 | 4 | 5 | 6 | 7 | 8 | 9 |
| --- | --- | --- | --- | --- | --- | --- | --- | --- | --- | --- |
| Depression severity | Group |  |  |  |  |  |  |  |  |  |
| **Minimal** | *bd* | 0 | 0 | 0 | 0 | 0 | 0 | 0 | 0 | 0 |
|  | *bpd* | 0 | 0 | 1 | 1 | 0 | 0 | 0 | 0 | 0 |
|  | *mdd* | 0 | 0 | 0 | 0 | 0 | 0 | 0 | 0 | 0 |
| **Mild** | *bd* | 1 | 1 | 1 | 1 | 1 | 1 | 1 | 0 | 0 |
|  | *bpd* | 0 | 1 | 1 | 1 | 1 | 1 | 1 | 0 | 0 |
|  | *mdd* | 1 | 1 | 1 | 1 | 1 | 1 | 1 | 0 | 0 |
| **Moderate** | ***bd*** | 1 | 1 | 2 | 2 | 2 | 2 | 2 | 0 | 1 |
|  | ***bpd*** | 1 | 1 | 2 | 1 | 1 | 2 | 2 | 0 | 1 |
|  | ***mdd*** | 1 | 1 | 2 | 2 | 1 | 2 | 1 | 0 | 0 |
| **Moderately Severe** | ***bd*** | 1 | 2 | 3 | 2 | 3 | 2 | 2 | 1 | 2 |
|  | ***bpd*** | 2 | 2 | 2 | 3 | 2 | 3 | 2 | 0 | 1 |
|  | ***mdd*** | 2 | 2 | 3 | 3 | 2 | 3 | 2 | 1 | 1 |
| **Severe** | ***bd*** | 3 | 3 | 3 | 3 | 3 | 3 | 2 | 2 | 3 |
|  | ***bpd*** | 3 | 3 | 3 | 3 | 3 | 3 | 3 | 1 | 2 |
|  | ***mdd*** | 3 | 3 | 3 | 3 | 2 | 3 | 3 | 1 | 3 |

**Table S3:** Nine-item patient health questionnaire (PHQ-9) item descriptions, as introduced by Kroenke and Spitzer (2002).

| **PHQ-9 item no.** | **Description** |
| --- | --- |
| **1.** | Little interest or pleasure in doing things |
| **2.** | Feeling down, depressed, or hopeless |
| **3.** | Trouble falling or staying asleep, or sleeping too much |
| **4.** | Feeling tired or having little energy |
| **5.** | Poor appetite or overeating |
| **6.** | Feeling bad about yourself — or that you are a failure or have let yourself or your family down |
| **7.** | Trouble concentrating on things, such as reading the newspaper or watching television |
| **8.** | Moving or speaking so slowly that other people could have noticed? Or the opposite — being so fidgety or restless that you have been moving around a lot more than usual |
| **9.** | Thoughts that you would be better off dead or of hurting yourself in some way |

**Figure S1.**
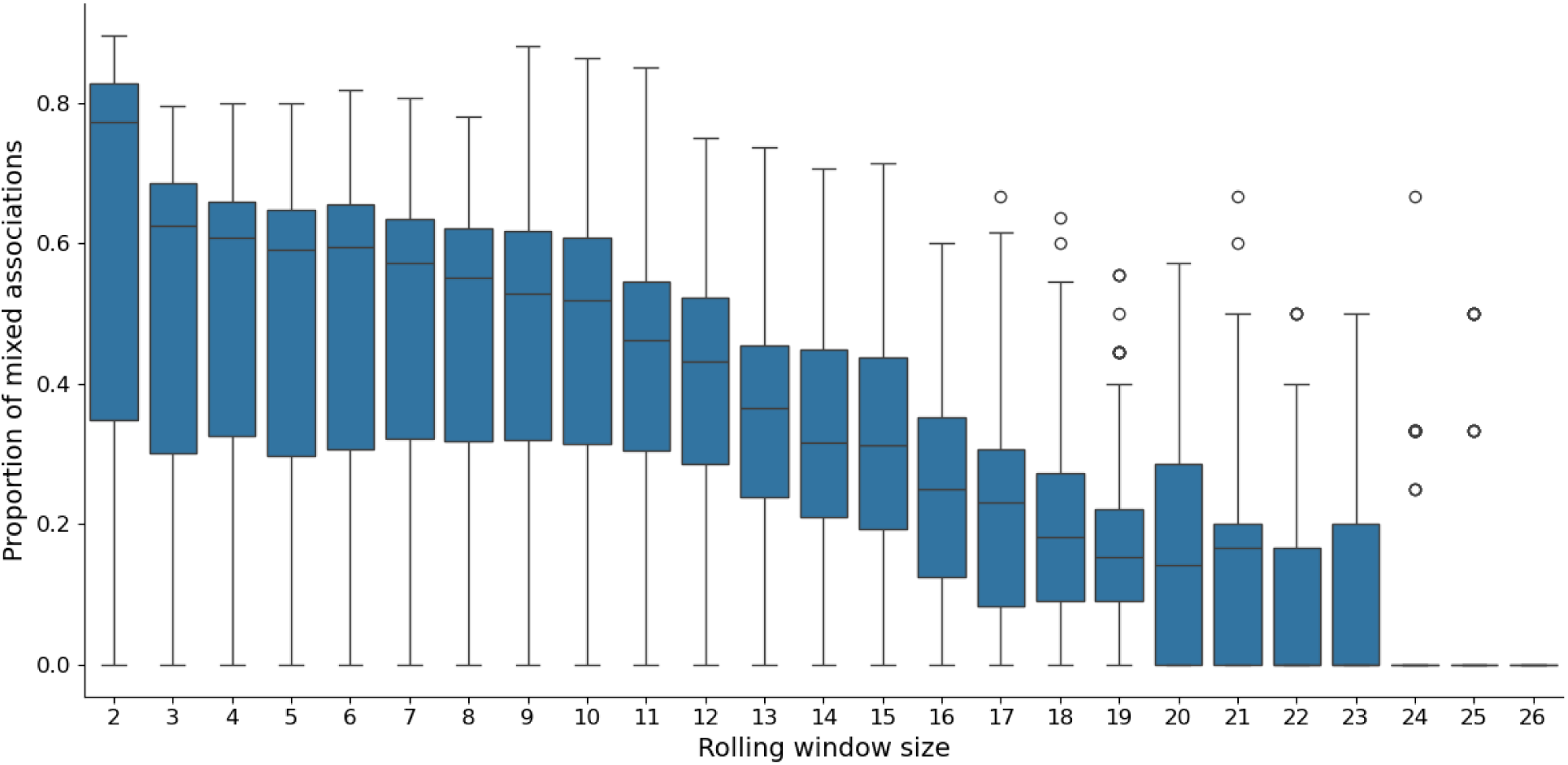
Exploratory rolling window correlation analysis summary, showing the proportion of behavioral features showing both positive and negative Kendall’s Tau rank correlation coefficients across the users. In general, shorter window yields higher proportion of mixed correlations. Window size of 2 (equaling 28 days) result in 67% mixed correlations on average.

**Table S4.**
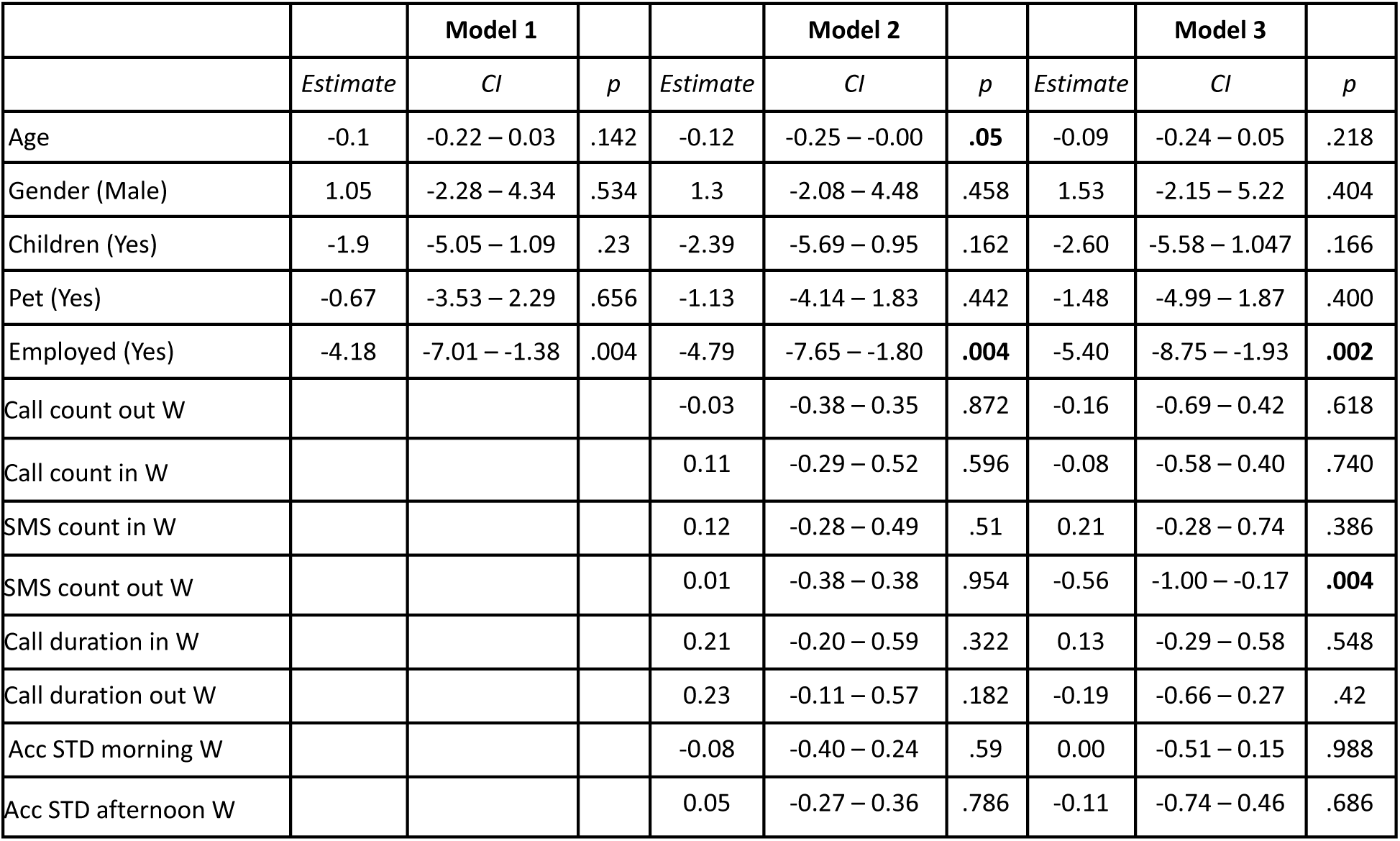
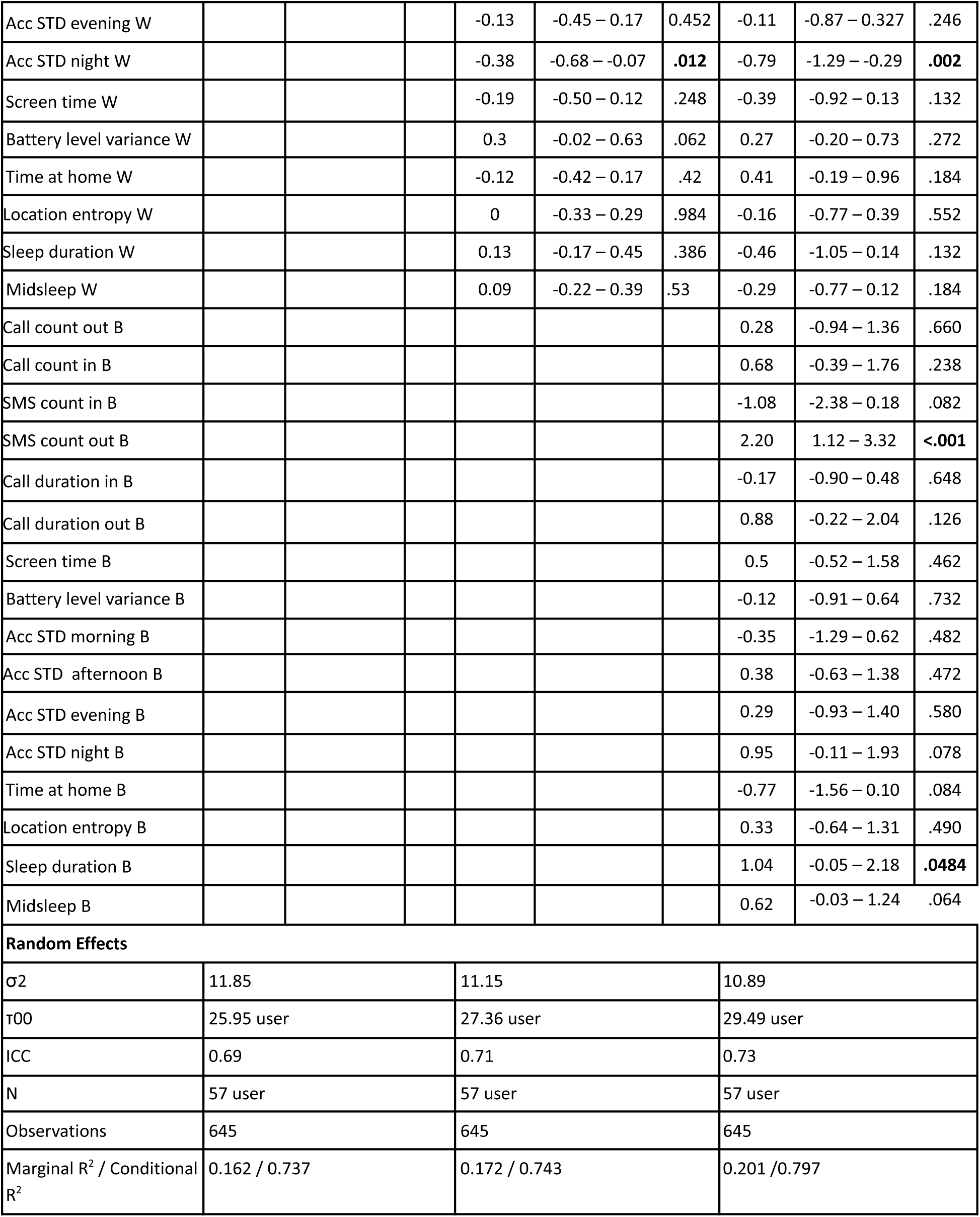
Multilevel model summaries. Model 2 is the best fitting and parsimonious, showing within-participant behavioral features: (1) daily outgoing SMS count associated higher depression severity, and (2) nighttime accelerometer variability with lower severity. Additionally, higher age and employed status is associated with lower depression severity. Notably, between-participant effects did not show any statistically significant associations. W denotes within-participant effect, B between-participant effect, σ2 residual variance, τ00 intercept variance, and ICC intra class correlation coefficient. Model 1 includes background demographics and contextual predictors, with random intercepts for each participant. Model 2 adds within-particiant effects (features) into model 1 Model 3 adds between-participant effects (features) into model 2.

## Multimedia Appendix 2

**Table S1:** Summary of extracted features across different behavioral aspects, including physical activity, mobility, sociability, phone interactions, and sleep. Features are derived from smartphone sensor data and logs; accelerometer, GPS, phone call and SMS logs, application usage, and battery levels. Sleep features were inferred from screen usage data.

| Aspect | Sensor | Raw features |
| --- | --- | --- |
| Physical activity | Accelerometer | Acceleration magnitude, magnitude max, min, mean and standard deviation, measurement count |
| Mobility | GPS location | *Distance-based:* total distance, variance, log variance, average speed, speed variance, max speed, location bin count.<br>*Significant place-related:* static point count, moving point count, static bin count, max distance from home, number of significant places, number of rarely visited places, number of transitions between significant places, bin count in top1, top2, top3, top4, and top5 cluster, normalized entropy |
| Sociability | Phone call and SMS logs | *Phone call-related:* Total call duration, call duration mean, median and standard deviation, call count, type (incoming and outgoing)<br>*SMS-related:* SMS count, type (incoming and outgoing) |
| Phone interactions | Application and screen usage, battery charge level | *Application-related:* Application class, count, duration<br>*Screen usage-related:* screen-off timestamp, screen event count, screen event duration, screen event duration max, mean, min, and standard deviation, screen first unlock timestamp<br>*Battery level related:* Battery level mean, median and standard deviation, battery shutdown time, battery discharge |
| Sleep | Inferred from screen | Average sleep duration, sleep start time, sleep end |
|  | usage data | time, midsleep time |

## Multimedia Appendix 3

Variables for the multilevel model were selected to capture aspects of behaviors based on prior evidence linking behavioral markers—such as sleep, activity rhythms, communication patterns, mobility, and device usage—with depressive symptoms. Sleep: duration and midsleep point. Temporal activity rhythm: standard deviation of magnitude from the accelerometer for four segments of days. Communication: count of ongoing and incoming sms/calls, duration of outgoing and incoming sms/calls. Location: time at home and entropy of location. Device usage: screen usage duration, battery mean level. The model specification is presented below.

PHQ9-score ∼ Age + Gender + Children + Pet + Employed +

OutgoingCallsCount.between + OutgoingCallsCount.within +

IncomingCallsCount.between + IncomingCallsCount.within +

OutgoingSMSCount.between + OutgoingSMSCount.within +

IncomingSMSCount.between + IncomingSMSCount.within +

OutgoingCallDuration.between + OutgoingCallDuration.within +

IncomingCallDuration.between + IncomingCallDuration.within +

ScreenTime.between + ScreenTime.within +

BatteryVar.between + BatteryVar.within +

AccelMorning.between + AccelMorning.within +

AccelAfternoon.between + AccelAfternoon.within +

AccelEvening.between + AccelEvening.within +

AccelNight.between + AccelNight.within +

TimeAtHome.between + TimeAtHome.within +

LocationEntropy.between + LocationEntropy.within +

SleepDuration.between + SleepDuration.within +

Midsleep.between + Midsleep.within +

(1|pid)

